# Translation of clinical practice guidelines on lifestyle behavior in a psychiatric setting. A discourse analytical case study

**DOI:** 10.1101/2024.01.18.24301407

**Authors:** Cathrine Fonnesbech Hjorth, Louise Lund Thomsen, Jan Mainz, Henrik Bøggild

**Affiliations:** Public Health and Epidemiology Group, Department of Health Science and Technology, Aalborg University, Denmark; Psychiatry Management, North Denmark Region, Aalborg, Denmark; Department of Community Mental Health, Haifa University, Israel; Department for Health Economics, University of Southern Denmark, Denmark; Danish Center for Clinical Health Services Research, Department of Clinical Medicine, Aalborg University, Aalborg, Denmark

**Keywords:** Translation, Discourse analysis, Front-line nurses, Lifestyle, Psychiatry, clinical practice guidelines

## Abstract

**Introduction:** Implementation of clinical practice guidelines (CPGs) can be constrained by diverging guideline interpretations, leading to a discrepancy between the intended clinical practice and the clinical practice implemented. We examine how CPGs on lifestyle behavior for psychiatric inpatients are translated into clinical daily practice, and how this was affected by discourses surrounding the stakeholders.

**Methods:** We conducted a comparative case study within two psychiatric wards in Denmark, wherein a clinical practice guideline (CPG) addressing lifestyle behavior was implemented. The objective of the CPG was to strategically intervene in the prevalence of unhealthy lifestyle practices among inpatients. Critical discourse analysis and a perspective of translation formed the theoretical framework. We collected empirical material at two stakeholder levels; the CPG authors and clinicians. This included documents related to the CPGs, field observations and two focus group interviews with clinicians.

**Results:** The CPG were composed within a system-centered discourse where lifestyle intervention was considered relevant for all inpatients. The clinicians translated the CPGs within a person-centered discourse and a prioritization discourse, focusing on individual factors such as the patient’s own wishes, surplus energy, and the status of the psychiatric treatment.

**Conclusion:** The findings show that implementation of guidelines in their original form cannot be anticipated, but will constitute translation based on social structures and their discourses. Unawareness of this phenomenon called translation may affect implementation and should be considered when CPGs are developed and launched. Future research should explore how discourses and social processes among patients influences implementation of CPGs.

**What is known on this topic?:**

- Several barriers explaining why clinical practice guidelines are not followed by intention has been reported
- Less is known about the situations in which the clinicians believe that they follow the guidelines, but translate the content of them unintentionally

**What this study adds to existing knowledge:**

- Our findings show how clinical practice guidelines can be translated according to the discourses and social processes surrounding the different stakeholders. In this study, clinicians unintentionally changed the practice intented in the guidelines.
- The clinical practice guidelines arose from a system-centered discourse, expecting all patients to have the same need for lifestyle intervention.
- The dissemination of the clinical practice guidelines was formed by a patient-centered and a prioritations discourse, focusing on the patients’ individual needs.

**What are the implications for practice?:**

- Clinical practice guidelines are translated differently by stakeholders, and cannot be expected to be implemented in its original form.
- Policy makers should be aware of this phenomenom called translation, as it is pivotal to align perceptions of clinical practice guidelines or interventions across stakeholders.

## Introduction

Manifested by increased mortality^1^ and somatic comorbidity,^2,3^ unhealthy lifestyle behavior such as unhealthy diet, alcohol abuse, inactivity and smoking, is common in people with psychiatric diseases. Therefore, a wide range of nutrition guidance, physical activity and smoking interventions emerge globally in psychiatric settings.^4–7^ Such efforts add to the traditional clinical practice at psychiatric wards, arguing for clinical practice guidelines (CPGs) when implementing lifestyle interventions. However, implementation of lifestyle CPGs can be problematic in a psychiatric setting as established practices can be challenging to change, and the benefit can be limited.^8,9^

CPGs are designed to assist clinicians in their daily clinical practice and as a tool to implement new interventions.^10^ A downside of CPGs is that they might interfere with clinical judgment.^11^ Implementation of CPGs is often inadequate, expressed by a discrepancy between the composed, written content and the practice carried out to the patients.^12–18^ CPGs not being implemented means that evidence-based practice in health services may be insufficient and recommended practices are not provided to the patients.^17^

CPGs are typically developed by academics and managers (hereafter CPG authors) as directive documents the clinicians are recommended, if not expected, to implement in clinical practice. Lack of motivation and consensus, and attitude towards the CPGs are some of the most frequently reported barriers towards implementation among clinicians.^19,20^ This might explain why CPGs are not followed intentionally, but less is known about the situations in which the clinicians *think* that they follow the guidelines, but translate the content of them unintentionally. This study examines the underlying processes leading to this phenomenon of translation in order to understand why CPGs are sometimes understood differently when adopted in clinical practice. We use a critical discouse analysis (CDA) to examine the ways CPGs are translated, aiming to unveil the discourses in which CPG authors’ production of the CPGs and the clinicians’ receiving of them take place.

## Theoretical framework

To gain an understanding of the social processes rooted in the discrepancy between the intention of the CPGs and actual clinical practice, we use Richard Freeman’s translation framework, aligned with CDA and Pierre Bourdieu’s social theory. Together, these frameworks and theories create a critical lens through which we explore not only *what* is happening, but also *why* and *how* it is happening.

### The translation perspective

Translation in the sense used here is concerned with how policy translates when moving from one place to another. With this focus, we distance ourselves from an orthodox way of seeing policy transfer as policy implementation in its original form. Freeman’s perspective considers translation as ongoing changes and negotiation processes by which policies are never considered fully developed. Instead, they are subject to continuous interpretation and representation by the different stakeholders interacting with the policies. ^21^ According to Freeman,^22^ an understanding of these changes and negotiation processes can be enhanced by exploring transitions that policies undergo from their production to their instantiation in daily practice:

> “To understand implementation, then, which we might think of as the realisation of documents in practice, we need to understand what happens in the spaces between them. How is one document translated into another, by whom and what for? How is one document articulated with and in another? Even to begin to answer such questions, we need to know how documents are written and read, produced and received” ^22^ (p. 162)

The translation of policy into different social structures depends on stakeholders and the social context in which the policy is implemented. Hence, implementation is evolutionary in the sense that the original policy will be reformulated when moving through stakeholders and contexts within the organization’s different hierarchal levels.^23^ As such, we see translation as a social phenomenon and advocate that an understanding of implementation demands insight into the social interactions surrounding the CPGs. Insight into *why* and *how* the texts are produced and received is examined using Norman Fairclough’s CDA.^24^

### Critical discourse analysis

CDA is a text-oriented, interdisciplinary approach combining linguistically oriented textual analysis with sociological principles.

The ontological and epistemological premise of CDA is that reality is created through our articulations and the added significance of the words in the contexts in which they are used. Thus, reality is a product of our spoken words and the position from which we speak.^24^ CDA can uncover how stakeholders draw on discourses in their articulation of the CPGs and how these articulations diverge within and between stakeholders across hierarchical levels in the healthcare system. Fairclough understands discourse as language used as a social practice. In other words, as a way of speaking from a certain perspective.^24^ Language, including its linguistic elements, thus helps construct the social world’s subjects, relationships, knowledge, meanings and their power relations. According to Fairclough, social practices can also be non-discursive. In this sense, the structure of the healthcare system can be perceived as a non-discursive practice that may affect how the clinicians prioritize CPGs.^24^ Therefore, we used CDA to examine how CPGs are composed and translated by discursive and non-discursive practices.

CDA can reveal how discursive practices help maintain unequal power relationships and thus suppression of social groups.^24^ In the present case, we will argue that unequal power relationships exist between the CPG authors and the clinicians, as the CPG authors have legitimated power to issue CPGs that must be followed by the clinicians. Additionally, politicians influence the practices of the CPG authors. Although we did not directly study the role of the politicians in this study, their position in the hierarchy of power relations will be included in the analysis. The hegemonic struggle between the discourses in the field is part of the larger social practice that contributes to the reproduction and transformation of the CPGs and thus their translation. The stakeholders’ resources, which is Fairclough’s collective term for knowledge, common sense and ideological conviction^24^ are included in the analysis of the power struggles to identify and explain dominant relationships. We included Bourdieu’s social theory to further examine these social practices.

### Bourdieu’s social theory

Bourdieu understands individuals as active social agents who relate to reality and the society they live in. Hence, it is from this position their actions are performed and their choices are made.^25^ Based on this perspective, different understandings of the CPGs and miscellaneous relations to the clinical setting are expected. According to Bourdieu, part of an individual’s conceptions of the world is non-verbal and embedded. To expound how this implicit knowledge contributes to the translation of the CPGs we use Bourdieu’s concepts habitus, capital and doxa. Capital, in Bourdieu’s understanding, can be translated as resources or skills of social practice underlying a person’s power and influence in a particular field. It occurs in three forms; *economic, cultural* and *social* capital. When capitals in any form are accepted and added value in a field they are considered a *symbolic* capital. Economic capital relates to money and material possessions.^26^ For instance, the economic capital of politicians and the CPG authors is institutionalized by their right to command and allocate financial resources. In the form of symbolic capital, this may give them a powerful position in the field. Cultural capital includes education, titles, licenses, and certificates.^26^ Clinicians have cultural capital by virtue of their experience and education, which encourage patient care, whereas the cultural capital of the CPG authors may favor an evidence-based, cost-effective health care system. Social capital is a resource in the form of network-based collaborations and social relationships.^26^ As emphasized later, the clinicians differentiated by being a group of healthy role models at one of the wards, and a group of clinicians who generally did not refer to themselves as such. The density of role models in a group will add value the social capital in the group and likely support a focus on healthy lifestyle. We expect that the capital of the different stakeholders related to the CPGs differ and add different value to their articulations and actions in the field.^26^ We use the concepts of capital to explain the positions of different stakeholders under the influence of certain discourses. Habitus is the bodily embedded systems of dispositions acquired within an environment and the habits, attitudes, and behaviors of the stakeholders.^25^ We use this notion to understand the stakeholders’ perceptions and articulation of the CPGs and to explain why the stakeholders construct them differently. Doxa covers what is taken for granted in a field; the common-sense assumptions about right and wrong, normal and deviant, qualified and disqualified. Doxa is embedded in the stakeholders’ habitus but is also part of the field.^27^ We assume that among the CPG authors a doxa is that clinicians act as anticipated in the CPGs.

The hierarchal position and power of stakeholders in a field are constituted by habitus and capital. Using these concepts contribute to the analysis of the dialectical relationship between discourses and non-discursive social practices and make it possible to describe the non-discursive practices because the concepts can help embrace the unsaid. Power and dominance relationships among the stakeholders can be described with the concept of symbolic violence. Violence is understood as a domination that arises in the interaction between stakeholders and is characterized by the unconscious acceptance of the dominated.^27^ This could be the politicians or CPG authors’ possession of economic capital, which puts them in a position to dominate over the clinicians.

## Methods

### Setting

In autumn 2015, we conducted a comparative case study with two psychiatric wards we name “ward A” and “ward B”.^28^ Both wards were implementing the same lifestyle CPG, enabling us to analyze possible causes for different translations influencing the implementation. The wards were both administratively part of the the Department of Psychiatry, North Denmark Region. The wards had the same ultimate superiors, but separate immediate managers and clinicians. We opted to examine two geographically distinct wards characterized by varying patterns in admission diagnoses amnd contexts, increasing the possibility of obtaining a valid generalization.^28^ Ward A provided care for people diagnosed with schizophrenia or other psychotic illnesses and participates in a Danish patient safety program for mental health, “Safe Psychiatry”, focusing on timely and efficient diagnosis and treatment of somatic comorbidities.^29^ Ward B provided care for people diagnosed with depression or bipolar disorders.

CPGs concerning lifestyle prevention initiatives had been a part of the expected clinical practice at the wards since 2006. The CPGs are developed according to national policies, and describe a lifestyle program consisting of 1) lifestyle screening at hospitalization and 2) an offer of in-hospital intervention targeted lifestyle risk factors identified in the screening or in general. The screening includes a clinical examination consisting of measures of body mass index and waist circumference, and interviews about smoking, nutrition and physical activity habits. The intervention consists of counselling, including motivational interviewing and possibly individual programs. Both wards offered weekly group sessions concerning prevention of lifestyle risk factors, outdoor walking activities and equipment for exercise. The implementation of the lifestyle program was assessed quantitatively using run charts.^30^ Fulfilment of the process indicators was not yet achieved at the wards (Supplementary File 1). Notably, the clinicians stated that they fulfilled the CPGs, suggesting that they had a different understanding of the content, than captured in the quantitative assessment.

### Empirical material

CDA is based upon spoken and written words representing discursive aspects of the CPG authors and clinicians. We link the texts to broader social practices to achieve an in-depth understanding of factors influencing the different perceptions of the CPGs ^24^. Figure 1 presents an overview of the empirical data generated. For the CPG authors’ perception of the content, data is based on the written CPGs and policy documents forming the basis of the CPGs. The statutory policy document *The Psychiatry Plan 2015-2018* ^31^ describes the visions of the psychiatric sector in the North Denmark Region. Two *action plans* describe the practice standard related to the CPGs, including information on how they should be performed and by whom at the specific wards. Finally, two national documents describing a Danish patient safety program *Safe Psychiatry*^29^ concerning lifestyle prevention strategies were included, as ward A participated in this program with relevance to their action plan and potentially their translation of the CPGs.

**Figure 1:**
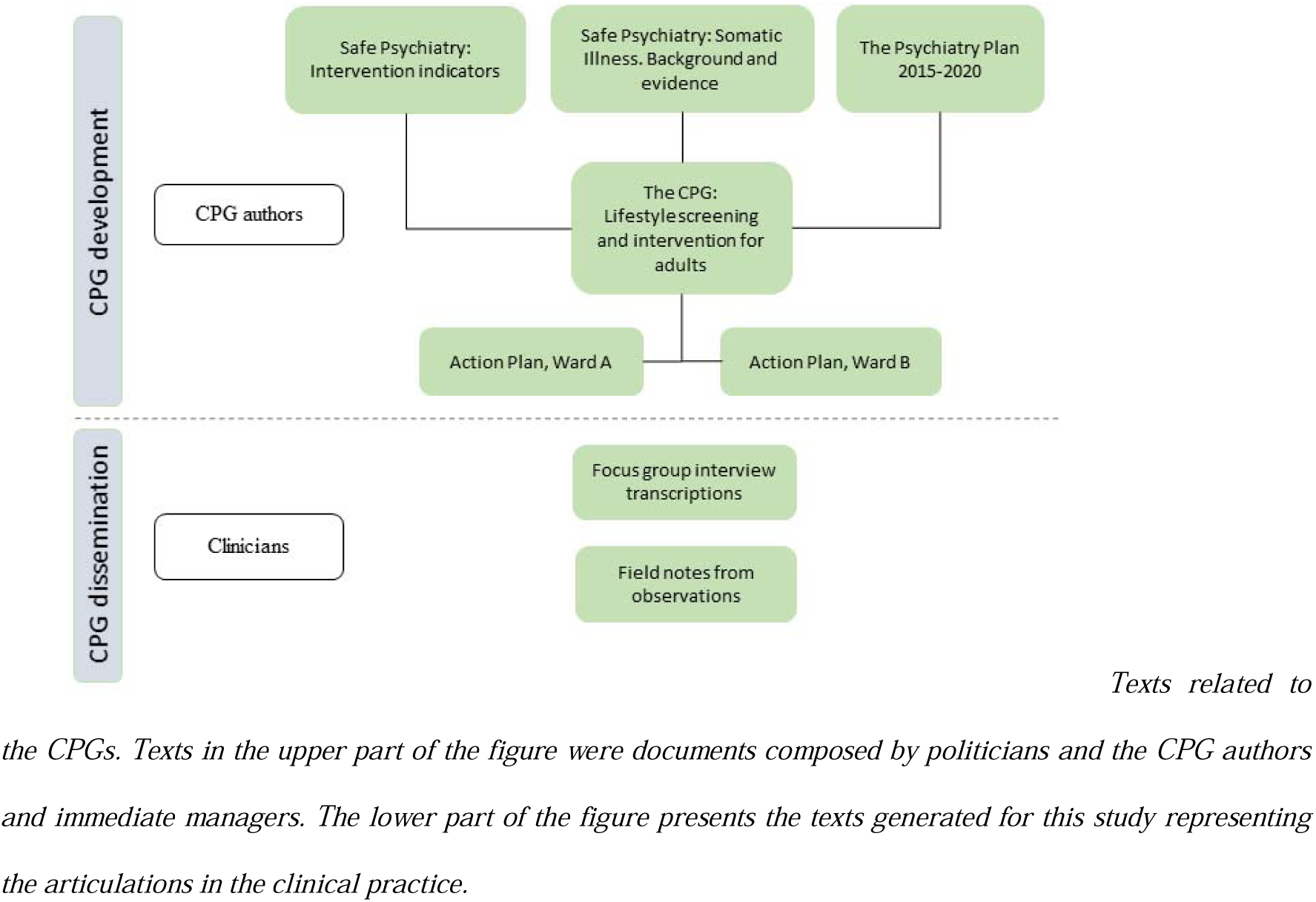
Empirical data included in the critical discourse analysis across stakeholder levels, North Denmark Region.

Clinicians’ perspectives were collected from field observations and focus group interviews by two researchers. Three field observations of one hour (two at ward A, one at ward B) were carried out. In addition, the researchers participated in-group sessions related to patients’ lifestyle behavior. Field notes were taken with the purpose to collect aspects of the daily-life articulations of the practices described in the CPGs. All healthcare workers at the wards were invited to participate in focus group interviews, regardless of their age, gender or seniority. Three nurses and two social and health care assistants participated in a focus group at ward A. A total of four nurses participated in a focus group at ward B. The focus group approach was selected specifically to allow interpersonal dialogue to take place, in order to analyze the articulation of the CPGs.

The focus groups interviews were semi-structured based on pre-determined open-ended questions and discussion points (Supplementary File 2). One researcher acted as moderator and handed out discussion cards, printed CPGs and the local action plans during the focus group interviews in order to facilitate a discussion of the clinicians’ perception of the CPGs. The other researcher noted down gesticulations and notable body language. The researchers aimed to establish a permissive environment and explained firmly that the purpose of the study was not to control whether the clinicians knew or followed the CPGs, but to learn how they experience and articulate the CPGs and lifestyle prevention. Focus group interviews were audio-recorded and transcribed into text by the researchers.

### Data analysis

The software program Nvivo version 10 was used as an analytical platform in the coding process and for providing an overview of the empirical evidence.

Fairclough developed a three-dimensional framework for studying discourse. The dimensions reflect three distinct but interrelated levels of the empirical analysis: 1) textual-linguistic analysis of spoken or written articulations, 2) analysis of discursive practice e.g. text production, distribution, and consumption and 3) analysis of discursive events. The textual dimension is rather descriptive, focusing on vocabulary, grammar and textual structures in the text.^24^ For instance, we analyze if stakeholders articulate with non-restricted or restricted clauses, indicating the positioning of the persons. The analysis of discursive social practices provides the link between text and social practice as it is in the discursive social practices that text is translated.^24^ In this part of the analysis, we explored how other texts were reproduced into new texts by analyzing intertextuality, the scenario where stakeholders articulate elements of other texts. This refers to interdiscursivity understood as occurrence and combination of discourses. The third dimension, the analysis of non-discursive social practices e.g. power struggles, sought to explain the discursive and non-discursive practices and their relationship to the broader social context.^24^

## Results

Three discourses were identified, their appearances in the empirical data are briefly summarized in Figure 2, and Figure 3 illustrates which among which stakeholders the discourses were identified.

**Figure 2:**
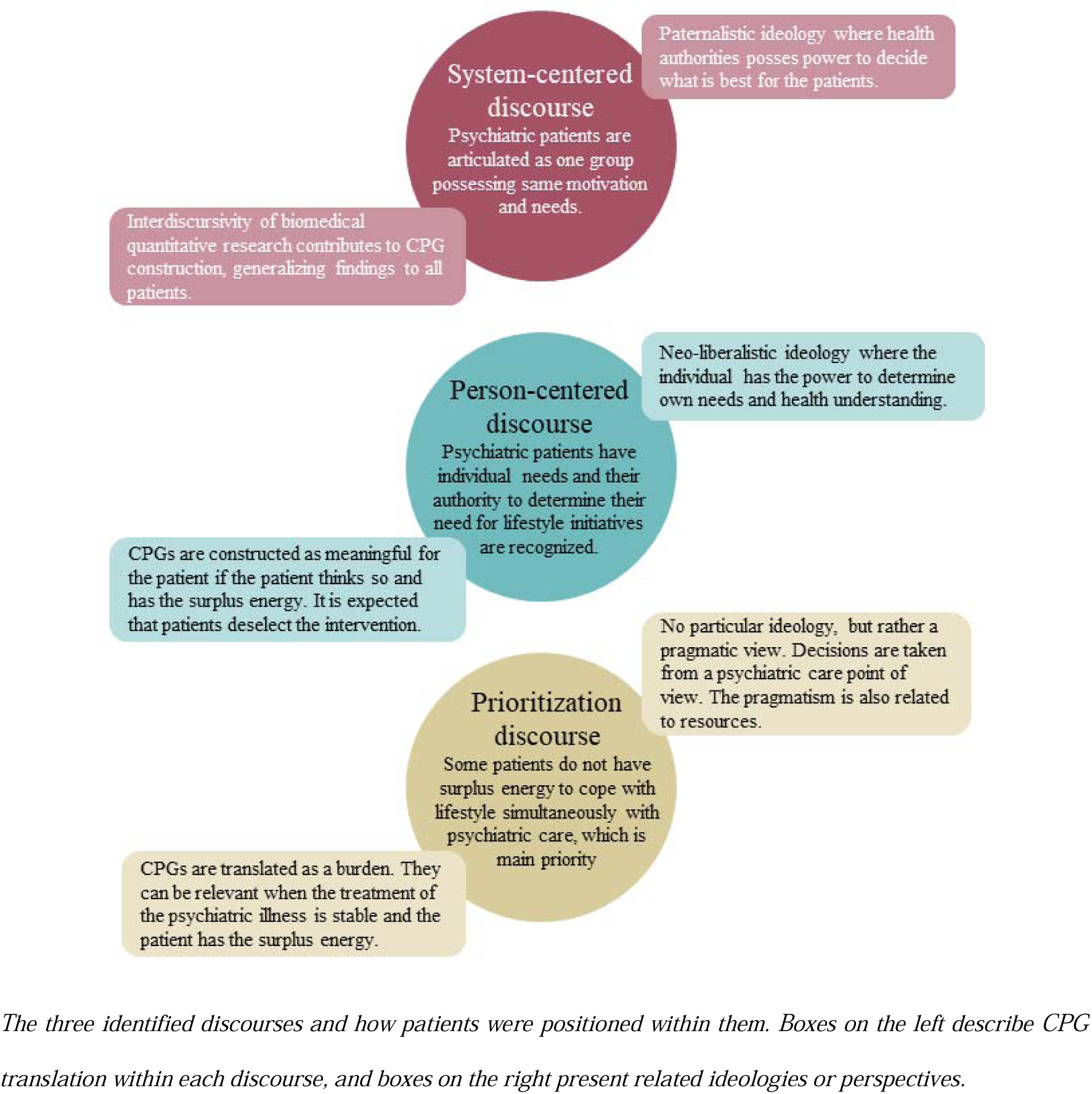
Characteristics of discourses dominating the construction and translation of the CPGs.

**Figure 3:**
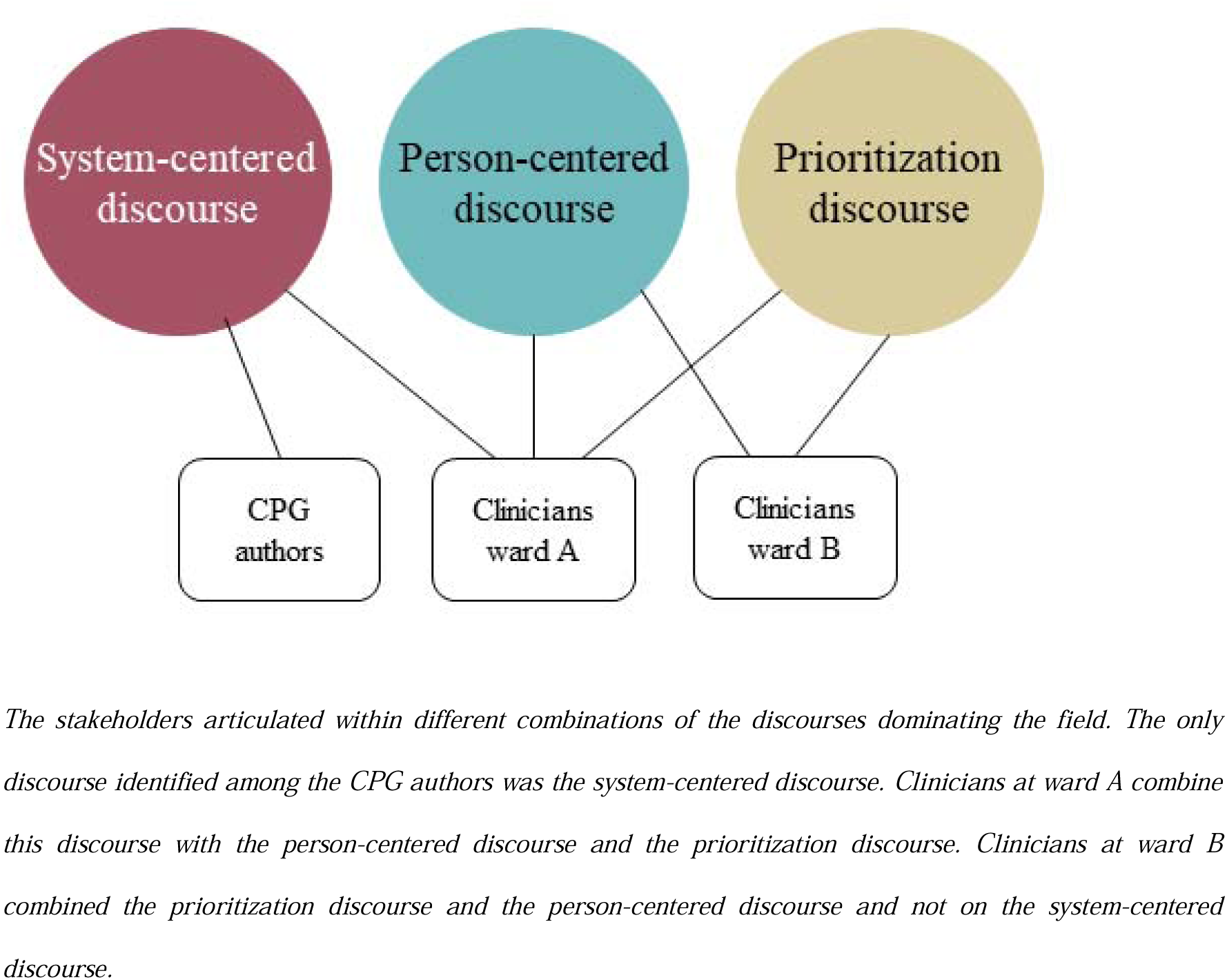
The discourses within the field and theit identification among stakeholders.

### The system-centered discourse

The system-centered discourse formed the documents composed by the CPG authors, but it was also identified among the clinicians at ward A in certain articulations. A dominant construction at this level is seeing all patients as having unhealthy lifestyle behavior, which denotes the system-centered approach embedded in this discourse.

The emphasis on language regarding psychiatric patients commonly treats them as a unified group, a perspective that may be perceived as doxa at this level of translation. The single set of CPGs, despite the diagnosis and patient characteristics, represents this doxa. The noun *people* (*e.g.: …with a mental illness)* is widely used in the policy documents and articulated in a nonrestrictive clause, implying that the statements regards all people with a mental illness. For instance, it appears as an assumption that all people with a mental illness are motivated to lifestyle changes and no attention is paid to potential different needs or motivations among people with severe mental illnesses. This was clear in the document *Safe Psychiatry.*^29^

> “Where there previously has been a perception that it was very difficult – if not impossible - for people with a mental illness to achieve lifestyle changes, we now know that these people are just as motivated as the rest of the population to achieve a healthier lifestyle”

The excerpt indicates a paternalistic ideology expecting everyone to desire a healthy lifestyle.^32^ Additionally, the patients are conferred low social and cultural capital, seen by the negative assertion *if not impossible*. This is translated in the CPGs, where lack of motivation is articulated as a problem for the clinicians to solve (hence not doxa):

> “A strengthening of the prevention at the hospital through early detection and recording of patient risk factors should support the psychiatric clinicians in the assessment of which patients are in need of a motivating, preventional and health promoting effort at the hospital, and which patients also need follow-up in general practice and optionally in their home municipality”.

Again, the CPG authors demonstrate a position of power to determine how patients should live.

The economic motive at the political level could be the driving force behind this, as the community stands to reduce expenses on care and treatment through the prevention of lifestyle diseases. The above excerpt also exemplifies an orthodox medical view, as the clinicians are given the power to assess patients’ needs, rather than acknowledging the patient’s own assessment.

Intertextuality to scientific research is common in the policy documents. This expert-oriented approach testifies a positivistic view. An example of this practice is seen in the document *Safe psychiatry,*^29^ where the system-centered discourse appears in the nonexistent distinguishing between diagnoses or illness severities when referring to scientific research:

> “On average, people with severe mental illness (including schizophrenia, bipolar disorder, schizoaffective disorder and severe depression) live 15-20 years shorter than the general population, and it is estimated that approximately 60% of this excess mortality is due to physical illness”.

The clinicians find the CPGs relevant for all patients, regardless of their lifestyle behavior. When the clinicians were asked what they thought the background of the CPGs was, one answered:

> “They come from the Danish Health Authorities, because psychiatric patients have excess mortality, and therefore, we have to do something about it” (Clinician, ward A)

The excerpt highlights the hierarchical structure and thus the power of the politicians and the clinicians’ acceptance of this. The high affinity of the clinicians’ use of the modal *must* express the staff’s duty in relation to the hierarchical order, which is perceived as legitimate.

In the CPGs and in the documents from the Safe Psychiatry the intervention part of the lifestyle program is described as an offer to the patient. However, the ward B action plan describes the performance of the preventive action passively as something *to work (…) with* and not as the clinicians’ duty. In the ward A action plan, the intervention appears as a duty taken for granted, and thus doxa, which in the clinicians’ articulations emerges as adapted. Thereby, the intervention part in the CPGs has been translated from being an offer to being mandatory at ward A. The action plan is composed locally within the ward contrary to the national documents Safe Psychiatry and the CPGs, and the translation may, therefore, be a product of the dialectic relationship between the discursive and non-discursive practices within the ward where the paternalistic ideology in the system-centered discourse seems partly adopted. Oppositely, the system-centered discourse was not identified at ward B. Later, we demonstrate that the clinicians’ habitus and capital across the wards varied, which presumably is important in this explanation.

### The person-centered discourse

The person-centered discourse reflected the autonomy and individual needs of the patients. The clinicians articulate a need to make person-centered assessments along with the patient and thus, the person-centered discourse was a contrast to the system-centered discourse. A clinician at ward B articulated personal resources as significant for the relevance of the lifestyle program:

> “Well, I think it depends on their, umm, on the environment they come from, their social background and their job situation. Actually, many of those we see have not had a job for many years and receive social security or something. This is actually of importance when they rate their own physical condition, compared to those who might have a good job and who are very conscious about it.

> Very athletic, exercise, eat healthy and eat….” (Clinician, ward B)

The language identifies the patients’ autonomy, profits, motivation and their own experiences of surplus energy and the clinicians are thereby moving towards a tradition of dialogue and caring. The articulation *they rate their own* exemplifies a view where the person’s own perception of their condition is applicable.

Resistance against the CPGs is particularly seen among clinicians at ward B, who lack a feeling of ownership of the lifestyle program, potentially due to dissociation from the paternalism in the system-centered discourse. Instead, individually targeted prevention appears as doxa. The clinicians at both wards manifest a lack of skills in individually targeted lifestyle prevention, illustrating the clinicians’ cultural capital and habitus being tied up around the psychiatric field and not to somatic diseases or lifestyle. Their expertise and skills are more aligned with the treatment of psychiatric illnesses rather than focusing on lifestyle prevention.. The below quotation exemplifies this, as a clinician at ward A requests a program more targeted the individual:

> “I could well imagine that we could improve the way it, now I speak for myself, but that we practiced more in individual prevention” (Clinician, ward A)

The incorporation of healthy lifestyle behavior into the clinicians’ habitus appears crucial for their acceptance and adherence to the CPGs.The clinicians question whether lifestyle prevention is a part of their profession and some feel uncomfortable about this enforced role. Unlike the clinicians at ward A, clinicians at ward B position themselves as *‘in the same boat’* as the patients when considering lifestyle behavior, opposing their ability to motivate patients to participate in lifestyle prevention. The clinicians at ward B translate the CPGs as an offer instead of a mandatory task. This seemed to be a translation affected by their habitus and cultural capital not accommodating a position as role models. Clinicians at ward A stand out as individuals in a field where a healthy lifestyle is dispositions of their habitus and where healthy lifestyle behavior as a goal itself in a paternalistic sense appeared as doxa. They focused on lifestyle interventions in the daily clinical practice and accepted the lifestyle program as a part of their profession.

### The prioritization discourse

The prioritization discourse is a pragmatic and ethical discourse counterbalancing the psychiatric care and the lifestyle program. This discourse appears primarily in the clinicians’ articulations. In the policy document and the CPGs, no precautions are taken against the question of priority of psychiatric care versus the lifestyle program. As seen above, the lifestyle program are targeted all psychiatric hospitalizations. The clinicians, however, stress the necessity to adjust the practice to the patient, as they consider psychiatric care as the main priority:

> “I cannot help thinking… when I think of this priority of prevention, I think okay, it is the disease that is the focus! In a way, first … We need to deal with this psychiatric diagnosis and focus on prevention in this regard. I do not think decidedly on lifestyle risk factors” (Clinician, ward B)

This exemplifies that the clinicians oppose the psychiatric care and the lifestyle program against each other in a matter of priorities, where the psychiatric care has priority exemplified through the expression *In a way, first*. That something needs to be handled *first* may both indicate that the clinicians need to prioritize their treatment because of limited resources and that the patient might lack the surplus to handle both. The lifestyle program still seems to be an effort provided when clinicians are aware of it and not as doxa at this level of translation. The clinicians at ward A experience that the lifestyle program sometimes outshines the psychiatric focus:

> “We have had to implement in relation to this lifestyle… And right now, we have entered a period where we have to find a balance, because…. it must of course not predominate everything else, but it is just as important as the patient’s psychiatric state. Because, as we worked a lot to make these care plans relative to the lifestyle program, right? It almost ended with, eh, lifestyle factors were the only thing mentioned in the care plans” (Clinician, ward A).

The opening *we have had to* expresses lack of ownership of the lifestyle program and the articulation *to this lifestyle* reflects the clinicians outdistancing the CPGs, indicating that it is not an internalized part of the clinicians’ profession and hence, their professional habitus. The conversation implies that the psychiatric care and the lifestyle program should be equally prioritized, or at least the psychiatric care should have the prime focus.

Several clinicians articulate that many patients do not have the *surplus energy* to work with lifestyle behavior during their hospitalization, because of their psychiatric condition. This makes the clinicians state prioritization needs. They use the object *surplus* repeatedly during the focus group interviews as something that legitimizes declination of participation in lifestyle interventions. The legitimization is emphasized in the following excerpt, where the clinician through the modality *well* legitimizes that inpatients deselect the intervention. The clinician express that the group sessions may be a legal excuse in order to fulfil the guidelines anyway:

> “Often…. Sometimes they decline [preventive conversations, ed.] because they lack the surplus energy and then I think: ‘well, the other [Group sessions, ed.] is mandatory to participate in… There, they have to participate” (Clinician, ward B).

The excerpt indicates that the clinician doubts that hospitalization is the appropriate context for lifestyle prevention. The negative assertion that the inpatients *lack the surplus energy* indicates that the clinician questions the relevance of the CPGs because the inpatients need energy to deal with their psychiatric treatment primarily.

The prioritization discourse appears as both constituents to and as constituted by the treatment of the psychiatric disease because this is more integrated into the clinicians’ professional habitus compared to the lifestyle program. This seems natural, as the psychiatric disease is the cause of the hospitalization and that the treatment of this is the clinicians’ primary task. Furthermore, the clinicians’ social practice can be constituted by non-discursive practice in terms of the Danish healthcare system where somatic and psychiatry are separate specialties and even hospitals.

## Discussion

The current study showed that surrounding discourses and underlying mechanisms which we have covered as habitus, capital forms and doxa highly influenced the translation of the CPGs and thus the implementation of them. The CPGs arose within a system-centered discourse, but were read in a field where the person-centered discourse and the prioritization discourse dominated. This entailed a gap between the CPGs and the actual daily practice. Discourses and the stakeholders’ habitus and capital forms did influence how the CPGs were translated and thus implemented, which we will elaborate in the following.

### Power struggles

Discursive power struggles are useful when explaining both the lack of consistency between the intended and the actual clinical practice, and why the clinicians at ward A translated the CPGs more alike the CPG authors than clinicians at ward B did. The person-centered discourse and the prioritization discourses were antagonizing the system-centered discourse and its position in the field of practice. The positioning of the individuals as one group in the system-centered discourse can be understood as a possible consequence of a surrounding evidence-based health care system, characterized by a positivistic tradition. The clinicians translated the guidelines from a neo-liberal ideology,^33^ contrary to the paternalistic ideology,^32^ thereby distancing from the evidence-based tradition. However, sometimes the system-centered discourse emerged among the clinicians at ward A, possibly because the intended practice matched their habitus and doxa. They copied some intentions of the CPGs and thus submitted to the CPG authors’ symbolic violence and reproduced the power relations between the CPG authors and the clinicians, though not without a struggle to find a balance between the physical life style program and the psychiatric care.

### Role models

It is a common experience that psychiatric nurses question lifestyle being part of the nursing role and that lack of skills and confidence can be a barrier for physical health care in a psychiatric setting.^8,34^ Freeman states that *”for what is written to become important, it must be talked about”.*^22^ It seems plausible that the lifestyle program is more interesting for the clinicians who act as role models, as this is accommodating their social capital in the group. Thereby, the habitus of the stakeholders affects how the documents are read collectively. This may explain why clinicians at ward A were more likely than clinicians at ward B to translate the CPGs into something mandatory and meaningful. This may also be affected by the structural, cultural and practical separation of the health care system, which has been identified earlier as a barrier for implementation of physical health care in a psychiatric setting.^34^

### Evidence-based practice

The capital of the CPG authors was tied up in an evidence-based practice, in contrast to the clinicians’ person centered stance and clinical judgment. Hodgson & Irving^35^ argue that politics in the last decades have become increasingly more evidence-based. They problematize a general understanding of evidence-based knowledge as an objective and rational truth.^35^ We found that the evidence-based approach did *not* outdo the clinicians’ role, as proposed by others.^11^ Instead, the consequence was the ‘cookbook’-approach were translated into what was considered meaningful in the clinical practice. The system-centered discourse may be constituted by discursive practices of a more general political level since politicians by virtue of their legislating power have a strong hierarchal position with the CPG authors following political decisions. Moreover, politicians wield economic power through the allocation of financial resources, thereby possessing significant economic capital, which grants them symbolic capital among the CPG authors. This way politicians may influence the discourses surrounding the CPG authors. Politicians are not typically equipped with the cultural capital necessary for evaluating evidence in healthcare. It appears that in the realm of politics, evidence is more a reflection of political alignment rather than a careful consideration of clinical practice contexts and patient needs. Conversely, clinicians may possess cultural capital to incorporate relevant evidence in their discursive practices. In line with this, Sackett et al.^36^ advocate that evidence-based ‘cookbooks’ should not replace clinical expertise and patients’ choices, but instead that clinical expertise should be used to judge the relevance of evidence in the particular setting.^36^

Quantitative assessment of the CPG is a practice within the system-centered discourse. The initial quantitative assessment documented a story of an implementation in progress, whereas our qualitative assessment told another story from the clinical practice, especially at ward B where few patients accepted the intervention and clinicians questioned the relevance. Comparable to this, a survey among physicians regarding their use of CPGs on cholesterol management revealed that 78% stated that they complied the guidelines, but interviews revealed that only 5% actually followed them.^37^ Obviously, this questions the usage of quantitative assessments in a field where documents travel between stakeholders. Our study extends the understanding of this phenomenon, and thus findings from other studies, by suggesting that the phenomenon can be explained by translation driven by discursive struggles.

CPGs often aim to reduce variations in practice,^13,20^ which seems paradoxical when clinicians and patients aim for more individualized treatment.^18,20^ Consequently, implementation is being limited by lack of flexibility in relation to individual needs and demands of the patients and lack of focus on the type of disease and severity.^13,38,39^ It is widely argued that acceptance and adherence of CPGs can be improved by customizing the guidelines by involving the end-users.^10,13^ Our study adds that the surrounding discourses should as well be investigated and integrated. Adding flexibility allowing space for clinical expertise and patients’ decisions when developing CPG on lifestyle behavior in psychiatric setting might support implementation.^10^ According to Freeman, translation is unavoidable,^21^ and its importance and influence was emphasized in our study. As such, social processes should be of interest when composing and implementing CPGs, probably also in settings other than the psychiatric wards, even discourses may differ. Inspiration on how to do this can be found in Kjell Arne Røviks work.^40^

### Limiation and strengths

CDA can be criticized for a weak theoretical understanding of social psychological processes, such as people’s control over their language in a given situation. Fairclough describes that discourses construct social identities and relationships, but a lack of empirical research on the processes limits the understanding of the stakeholder’s reproduction or change of discourse.^24^ We sought to overcome this by applying Freeman’s perspective of translation and Bourdieu’s sociological theory of difference to assist the CDA and provide a deeper understanding of the construction of the CPGs. The composition of Fairclough’s concepts in this study is based on the researchers’ preconceptions and thus a translation of Fairclough’s approach. Another composition of concepts and other preconceptions could possibly have led to different perceptions, discourses, and perspectives.

However, a transparent description of the methodological approaches to data collection and the analytical part is given explicitly and systematically. Thus, it is possible to assess whether the findings are transferable to other settings, if more similar casestudies across more psychiatric in-patient settings were undertaken. The systematic approach in the analysis reduced the impact of the preconceptions of the researchers and the theoretical perspectives contributed to distancing from the field, leading to new and potentially unconscious perspectives. To overcome abstract definitions of Fairclough’s concepts and analytic tools, we have presented our own understandings of the concepts and its application in order to make it transparent to the reader when concepts are applied. This has been a recurring reflection point in the research process and this transparency of subjective conceptual issues and critical attitude to own preconceptions helped to ensure validity in this study.^28^ We were not able to include the patients in this study, which could have extended the understanding of the translation of CPGs.

In summary, we experienced that implementation of CPG on lifestyle behavior in the two psychiatric wards relied on the translation of the CPGs. The translation happened through different discourses between involved stakeholders, which indicates that CPGs cannot be expected to be implemented as a structured approach in its original form, but will constitute and be constituted by social structures and their discourses. The stakeholders were unaware of the translation, and we recommend future initiatives to make this translation visible.

## Data Availability

All data produced in the present study are available upon reasonable request to the authors

## Acknowledgements

The authors thanks cand.scient.san.publ. Line Bilgrav, who participated in data collection and interpretation and acknowledge Clinic Psychiatry South in the North Denmark Region, for the valuable cooperation. A special thanks to the clinicians who participated in the focus groups, and to the patients who welcomed our presence at the wards.

## Ethics approval and consent to participate

The study was approved by the Psychiatry Management, North Denmark Region, as well as the ward managements. Patients and clinicians were given oral and written information about the study and were informed about researchers’ participation in group sessions and the aim of the study. All persons were allowed to decline the researchers’ attendance. The focus group participants were sent written information together with a consent form, which they signed before the interview.

