## Supplemental File 1 for "Translation of clinical practice guidelines on lifestyle behavior in a psychiatric setting. A discourse analytical case study"

Supplementary file 1 – Quantitative assessment of the implementation

### Quality assessment

As part of Safe Psychiatry, Clinic Psychiatry South continually evaluates the implementation process at ward A using process indicators. The process indicators in Safe Psychiatry measure "the proportion of discharges where the patient has received all elements of the Lifestyle program" using a composite indicator, where both screening and intervention must be documented. Moreover, the screening and the intervention are registered separately.^1^ We used the same approach to conduct a quantitative assessment at ward B. At ward A, the board of directors required screening and intervention of 95% of the inpatients cumulated. At ward B, the process indicator was divided into two, one monitoring the screening and one monitoring the intervention. Here, the goals were to screen 90%, and to intervene on 80% of the inpatients.

### Method

A run charts is a graphical depiction of the performance of process indicators, presented as time series that illustrate how the monitored initiative develops over time. With run charts and some simple rules it is possible to assess whether periodic outcomes for a specific time period are due to random variation or whether the variation is due to specific events at the given time.^2^ We used a reference table to assess variation, by comparing the number of eligible observations (those that do not fall on the median) with the following tools to assess variation: 1) the number of useful observations in the series diagram (observations equal to the median are not included), being the average for the performance of the indicator during the time period, 2) the number of times the curve crosses the median and 3) the number of observations that are above or below the median before the curve crosses again, here called the longest sequence.^3^ In addition to the number of usable observations, we used the reference table to determine a lower limit for the number of times the curve was allowed to cross the median and an upper limit for the longest series. If the median crossed less than one limit value or is the longest series longer than the other limit value, this was considered non-random variation, which is required when the effort is under development. It is only when the goal is reached that a random variation is desired. When this can be detected from a median that corresponds to the goal, it can be concluded that implementation is achieved and maintained.^2^ In the run charts below, the vertical axis shows percentages of discharges with received screening, intervention or both (composite graph). The horizontal axis shows the week numbers for the period (year 2015). The black curve represents fulfillment of the indicators over time, by interconnecting the observations. The quality goals are marked with a yellow horizontal line and the median for the period as a red horizontal line. Observations located on the median are not counted as useful observations in assessing the variation of the analysis. Only in case that a longer period of fulfilled indicators, which is not seen in the following analyses, a random variation is desired.

### Results

Assessment of medical records of all patients hospitalized for more 7 days or more during 2^nd^ March 2015-19^th^ October 2015 revealed that at ward A, screening was documented in 63% and intervention was documented in 29% of the inpatients. The cumulated indicator equaled 24%. However, the ward had improved remarkably in the second half of the period. At ward B, the cumulated indicator reached 57%. Screening was documented in 77% of the inpatients and intervention in 71% (however, at ward B declined intervention was documented as received. Restricting to actually received intervention, only 6% received the intervention). In conclusion, the run charts suggested that the lifestyle program was not implemented. The results are available below.

### Ward A

The run charts were conducted continuously by the administration at the Psychiatry in the North Denmark Region during the period, with 10 observations with minimum 14 days intervals. A total of 86 discharges were included.

**Figure S1. Implementation of the lifestyle program (both screening and intervention received)**

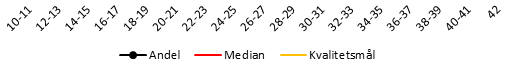

|  | Median | Mål | Andel alt eller intet |
| --- | --- | --- | --- |
| 2015-03-02 | 24.0909% | 95% | 0% |
| 2015-03-16 | 24.0909% | 95% | #N/A |
| 2015-03-30 | 24.0909% | 95% | #N/A |
| 2015-04-13 | 24.0909% | 95% | #N/A |
| 2015-04-27 | 24.0909% | 95% | 10% |
| 2015-05-11 | 24.0909% | 95% | 18.1818% |
| 2015-05-25 | 24.0909% | 95% | 0% |
| 2015-06-08 | 24.0909% | 95% | 30% |
| 2015-06-22 | 24.0909% | 95% | 16.6667% |
| 2015-07-06 | 24.0909% | 95% | #N/A |
| 2015-07-20 | 24.0909% | 95% | #N/A |
| 2015-08-03 | 24.0909% | 95% | #N/A |
| 2015-08-17 | 24.0909% | 95% | #N/A |
| 2015-08-31 | 24.0909% | 95% | 33.3333% |
| 2015-09-14 | 24.0909% | 95% | 37.5% |
| 2015-09-28 | 24.0909% | 95% | 85.7143% |
| 2015-10-12 | 24.0909% | 95% | 57.1429% |
| 2015-10-26 | 24.0909% | 95% | #N/A |
| 2015-11-09 | 24.0909% | 95% | #N/A |
| 2015-11-23 | 24.0909% | 95% | #N/A |
| 2015-12-07 | 24.0909% | 95% | #N/A |
| 2015-12-21 | 24.0909% | 95% | #N/A |
| 2016-01-04 | 24.0909% | 95% | #N/A |
| 2016-01-18 | 24.0909% | 95% | #N/A |
| 2016-02-01 | 24.0909% | 95% | #N/A |
| 2016-02-15 | 24.0909% | 95% | #N/A |
| 2016-02-29 | 24.0909% | 95% | #N/A |
| 2016-03-14 | 24.0909% | 95% | #N/A |
| 2016-03-28 | 24.0909% | 95% | #N/A |
| 2016-04-11 | 24.0909% | 95% | #N/A |
| 2016-04-25 | 24.0909% | 95% | #N/A |
| 2016-05-09 | 24.0909% | 95% | #N/A |
| 2016-05-23 | 24.0909% | 95% | #N/A |
| 2016-06-06 | 24.0909% | 95% | #N/A |
| 2016-06-20 | 24.0909% | 95% | #N/A |
| 2016-07-04 | 24.0909% | 95% | #N/A |
| 2016-07-18 | 24.0909% | 95% | #N/A |
| 2016-08-01 | 24.0909% | 95% | #N/A |
| 2016-08-15 | 24.0909% | 95% | #N/A |
| 2016-08-29 | 24.0909% | 95% | #N/A |
| 2016-09-12 | 24.0909% | 95% | #N/A |
| 2016-09-26 | 24.0909% | 95% | #N/A |
| 2016-10-10 | 24.0909% | 95% | #N/A |
| 2016-10-24 | 24.0909% | 95% | #N/A |
| 2016-11-07 | 24.0909% | 95% | #N/A |
| 2016-11-21 | 24.0909% | 95% | #N/A |
| 2016-12-05 | 24.0909% | 95% | #N/A |
| 2016-12-19 | 24.0909% | 95% | #N/A |
| 2017-01-02 | 24.0909% | 95% | #N/A |
| 2017-01-16 | 24.0909% | 95% | #N/A |
| 2017-01-30 | 24.0909% | 95% | #N/A |
| 2017-02-13 | 24.0909% | 95% | #N/A |
|  |  | 95% |  |
|  |  | 95% |  |
|  |  | 95% |  |
|  |  | 95% |  |
|  |  | 95% |  |
|  |  | 95% |  |
|  |  | 95% |  |
|  |  | 95% |  |
|  |  | 95% |  |
|  |  | 95% |  |
|  |  | 95% |  |
|  |  | 95% |  |
|  |  | 95% |  |
|  |  | 95% |  |
|  |  | 95% |  |
|  |  | 95% |  |
|  |  | 95% |  |
|  |  | 95% |  |
|  |  | 95% |  |
|  |  | 95% |  |
|  |  | 95% |  |
|  |  | 95% |  |
|  |  | 95% |  |
|  |  | 95% |  |
|  |  | 95% |  |
|  |  | 95% |  |
|  |  | 95% |  |
|  |  | 95% |  |
|  |  | 95% |  |
|  |  | 95% |  |
|  |  | 95% |  |
|  |  | 95% |  |
|  |  | 95% |  |
|  |  | 95% |  |
|  |  | 95% |  |
|  |  | 95% |  |
|  |  | 95% |  |
|  |  | 95% |  |
|  |  | 95% |  |
|  |  | 95% |  |
|  |  | 95% |  |
|  |  | 95% |  |
|  |  | 95% |  |
|  |  | 95% |  |
|  |  | 95% |  |
|  |  | 95% |  |
|  |  | 95% |  |

*The median fulfillment of the entire lifestyle program was 24.*

The composite indicator was not met during the period (Figure S1), but in progress illustrated by random variation, as the curve crosses the median more than two times and the longest sequence is <6 observations.

**Figure S2. Implementation of the screening part**

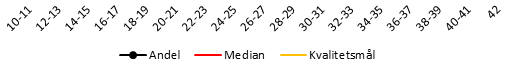

|  | median | mål | andel |
| --- | --- | --- | --- |
| 2015-03-02 | 63.3333% | 95% | 9.09091% |
| 2015-03-16 | 63.3333% | 95% | #N/A |
| 2015-03-30 | 63.3333% | 95% | #N/A |
| 2015-04-13 | 63.3333% | 95% | #N/A |
| 2015-04-27 | 63.3333% | 95% | 50% |
| 2015-05-11 | 63.3333% | 95% | 18.1818% |
| 2015-05-25 | 63.3333% | 95% | 42.8571% |
| 2015-06-08 | 63.3333% | 95% | 60% |
| 2015-06-22 | 63.3333% | 95% | 66.6667% |
| 2015-07-06 | 63.3333% | 95% | #N/A |
| 2015-07-20 | 63.3333% | 95% | #N/A |
| 2015-08-03 | 63.3333% | 95% | #N/A |
| 2015-08-17 | 63.3333% | 95% | #N/A |
| 2015-08-31 | 63.3333% | 95% | 66.6667% |
| 2015-09-14 | 63.3333% | 95% | 75% |
| 2015-09-28 | 63.3333% | 95% | 85.7143% |
| 2015-10-12 | 63.3333% | 95% | 100% |
| 2015-10-26 | 63.3333% | 95% | #N/A |
| 2015-11-09 | 63.3333% | 95% | #N/A |
| 2015-11-23 | 63.3333% | 95% | #N/A |
| 2015-12-07 | 63.3333% | 95% | #N/A |
| 2015-12-21 | 63.3333% | 95% | #N/A |
| 2016-01-04 | 63.3333% | 95% | #N/A |
| 2016-01-18 | 63.3333% | 95% | #N/A |
| 2016-02-01 | 63.3333% | 95% | #N/A |
| 2016-02-15 | 63.3333% | 95% | #N/A |
| 2016-02-29 | 63.3333% | 95% | #N/A |
| 2016-03-14 | 63.3333% | 95% | #N/A |
| 2016-03-28 | 63.3333% | 95% | #N/A |
| 2016-04-11 | 63.3333% | 95% | #N/A |
| 2016-04-25 | 63.3333% | 95% | #N/A |
| 2016-05-09 | 63.3333% | 95% | #N/A |
| 2016-05-23 | 63.3333% | 95% | #N/A |
| 2016-06-06 | 63.3333% | 95% | #N/A |
| 2016-06-20 | 63.3333% | 95% | #N/A |
| 2016-07-04 | 63.3333% | 95% | #N/A |
| 2016-07-18 | 63.3333% | 95% | #N/A |
| 2016-08-01 | 63.3333% | 95% | #N/A |
| 2016-08-15 | 63.3333% | 95% | #N/A |
| 2016-08-29 | 63.3333% | 95% | #N/A |
| 2016-09-12 | 63.3333% | 95% | #N/A |
| 2016-09-26 | 63.3333% | 95% | #N/A |
| 2016-10-10 | 63.3333% | 95% | #N/A |
| 2016-10-24 | 63.3333% | 95% | #N/A |
| 2016-11-07 | 63.3333% | 95% | #N/A |
| 2016-11-21 | 63.3333% | 95% | #N/A |
| 2016-12-05 | 63.3333% | 95% | #N/A |
| 2016-12-19 | 63.3333% | 95% | #N/A |
| 2017-01-02 | 63.3333% | 95% | #N/A |
| 2017-01-16 | 63.3333% | 95% | #N/A |
| 2017-01-30 | 63.3333% | 95% | #N/A |
| 2017-02-13 | 63.3333% | 95% | #N/A |
|  |  | 95% |  |
|  |  | 95% |  |
|  |  | 95% |  |
|  |  | 95% |  |
|  |  | 95% |  |
|  |  | 95% |  |
|  |  | 95% |  |
|  |  | 95% |  |
|  |  | 95% |  |
|  |  | 95% |  |
|  |  | 95% |  |
|  |  | 95% |  |
|  |  | 95% |  |
|  |  | 95% |  |
|  |  | 95% |  |
|  |  | 95% |  |
|  |  | 95% |  |

*The median fulfillment of the screening indicator was 63%.*

The increase in the fulfillment of the screening part (Figure S2) tends to be non-random variation, as the curve crosses the median only once, but more observations are needed to conclude this.

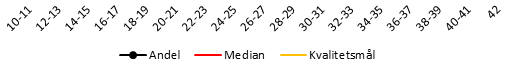
**Figure S3. Implementation of intervention part**

|  | Median | Mål | andel |
| --- | --- | --- | --- |
| 2015-03-02 | 28.6364% | 95% | 0% |
| 2015-03-16 | 28.6364% | 95% | #N/A |
| 2015-03-30 | 28.6364% | 95% | #N/A |
| 2015-04-13 | 28.6364% | 95% | #N/A |
| 2015-04-27 | 28.6364% | 95% | 10% |
| 2015-05-11 | 28.6364% | 95% | 27.2727% |
| 2015-05-25 | 28.6364% | 95% | 0% |
| 2015-06-08 | 28.6364% | 95% | 30% |
| 2015-06-22 | 28.6364% | 95% | 25% |
| 2015-07-06 | 28.6364% | 95% | #N/A |
| 2015-07-20 | 28.6364% | 95% | #N/A |
| 2015-08-03 | 28.6364% | 95% | #N/A |
| 2015-08-17 | 28.6364% | 95% | #N/A |
| 2015-08-31 | 28.6364% | 95% | 33.3333% |
| 2015-09-14 | 28.6364% | 95% | 50% |
| 2015-09-28 | 28.6364% | 95% | 85.7143% |
| 2015-10-12 | 28.6364% | 95% | 57.1429% |
| 2015-10-26 | 28.6364% | 95% | #N/A |
| 2015-11-09 | 28.6364% | 95% | #N/A |
| 2015-11-23 | 28.6364% | 95% | #N/A |
| 2015-12-07 | 28.6364% | 95% | #N/A |
| 2015-12-21 | 28.6364% | 95% | #N/A |
| 2016-01-04 | 28.6364% | 95% | #N/A |
| 2016-01-18 | 28.6364% | 95% | #N/A |
| 2016-02-01 | 28.6364% | 95% | #N/A |
| 2016-02-15 | 28.6364% | 95% | #N/A |
| 2016-02-29 | 28.6364% | 95% | #N/A |
| 2016-03-14 | 28.6364% | 95% | #N/A |
| 2016-03-28 | 28.6364% | 95% | #N/A |
| 2016-04-11 | 28.6364% | 95% | #N/A |
| 2016-04-25 | 28.6364% | 95% | #N/A |
| 2016-05-09 | 28.6364% | 95% | #N/A |
| 2016-05-23 | 28.6364% | 95% | #N/A |
| 2016-06-06 | 28.6364% | 95% | #N/A |
| 2016-06-20 | 28.6364% | 95% | #N/A |
| 2016-07-04 | 28.6364% | 95% | #N/A |
| 2016-07-18 | 28.6364% | 95% | #N/A |
| 2016-08-01 | 28.6364% | 95% | #N/A |
| 2016-08-15 | 28.6364% | 95% | #N/A |
| 2016-08-29 | 28.6364% | 95% | #N/A |
| 2016-09-12 | 28.6364% | 95% | #N/A |
| 2016-09-26 | 28.6364% | 95% | #N/A |
| 2016-10-10 | 28.6364% | 95% | #N/A |
| 2016-10-24 | 28.6364% | 95% | #N/A |
| 2016-11-07 | 28.6364% | 95% | #N/A |
| 2016-11-21 | 28.6364% | 95% | #N/A |
| 2016-12-05 | 28.6364% | 95% | #N/A |
| 2016-12-19 | 28.6364% | 95% | #N/A |
| 2017-01-02 | 28.6364% | 95% | #N/A |
| 2017-01-16 | 28.6364% | 95% | #N/A |
| 2017-01-30 | 28.6364% | 95% | #N/A |
| 2017-02-13 | 28.6364% | 95% | #N/A |
|  |  | 95% |  |
|  |  | 95% |  |
|  |  | 95% |  |
|  |  | 95% |  |
|  |  | 95% |  |
|  |  | 95% |  |
|  |  | 95% |  |
|  |  | 95% |  |
|  |  | 95% |  |
|  |  | 95% |  |
|  |  | 95% |  |
|  |  | 95% |  |
|  |  | 95% |  |
|  |  | 95% |  |
|  |  | 95% |  |
|  |  | 95% |  |
|  |  | 95% |  |

*The median fulfillment of the intervention indicator was 29%*

The intervention indicator was not reached during the period (Figure S3). However, it was in progress, illustrated by random variation, seen by the curve crossing the median twice and by the longest series of four observations.

### Ward B

The run charts were conducted by the researchers, with 10 observations with minimum 14 days intervals. A total of 93 discharges were included.

**Figure S4. Implementation of the lifestyle program (both screening and intervention received)**

|  | Andel | Median |
| --- | --- | --- |
| 10-11 | 66.6667% | 57.4809% |
| 12-13 | 60% | 57.4809% |
| 14-15 | 50% | 57.4809% |
| 16-17 | 33.3333% | 57.4809% |
| 18-19 | 57.1429% | 57.4809% |
| 20-21 | 40% | 57.4809% |
| 22-23 | 14.2857% | 57.4809% |
| 24-25 | 100% | 57.4809% |
| 26-27 | 40% | 57.4809% |
| 28-29 | 81.8182% | 57.4809% |
| 30-31 | 75% | 57.4809% |
| 32-33 | 50% | 57.4809% |
| 34-35 | 33.3333% | 57.4809% |
| 36-37 | 71.4286% | 57.4809% |
| 38-39 | 87.5% | 57.4809% |
| 40-41 | 66.6667% | 57.4809% |
| 42 | 50% | 57.4809% |

*The median fulfillment of the entire lifestyle program was 57%..*

Ward S7 did not state a quality goal for the composite indicator. The fulfillment of the entire lifestyle program (including both offered or received intervention), varied in the period (Figure S4). The variation is random, as the curve crosses the median seven times and the longest sequence of valid observations equals three.

**Figure S5. Implementation of the screening part**

|  | Andel | Median | Kvalitetsmål |
| --- | --- | --- | --- |
| 10-11 | 66.6667% | 76.5143% | 90% |
| 12-13 | 90% | 76.5143% | 90% |
| 14-15 | 100% | 76.5143% | 90% |
| 16-17 | 100% | 76.5143% | 90% |
| 18-19 | 77.7778% | 76.5143% | 90% |
| 20-21 | 80% | 76.5143% | 90% |
| 22-23 | 28.5714% | 76.5143% | 90% |
| 24-25 | 100% | 76.5143% | 90% |
| 26-27 | 60% | 76.5143% | 90% |
| 28-29 | 72.7273% | 76.5143% | 90% |
| 30-31 | 75% | 76.5143% | 90% |
| 32-33 | 50% | 76.5143% | 90% |
| 34-35 | 66.6667% | 76.5143% | 90% |
| 36-37 | 100% | 76.5143% | 90% |
| 38-39 | 100% | 76.5143% | 90% |
| 40-41 | 83.3333% | 76.5143% | 90% |
| 42 | 50% | 76.5143% | 90% |

*The median fulfillment of the screening indicator was 77%.*

The screening indicator was reached three times. The variation is random, as the curve with 17 usable observations crosses the median six times and since the longest series is shorter than five observations.

**Figure S6. Implementation of intervention part (both offered and received)**

|  | Andel | Median | Kvalitetsmål |
| --- | --- | --- | --- |
| 10-11 | 100% | 70.5672% | 80% |
| 12-13 | 62.5% | 70.5672% | 80% |
| 14-15 | 100% | 70.5672% | 80% |
| 16-17 | 33.3333% | 70.5672% | 80% |
| 18-19 | 57.1429% | 70.5672% | 80% |
| 20-21 | 50% | 70.5672% | 80% |
| 22-23 | 0% | 70.5672% | 80% |
| 24-25 | 100% | 70.5672% | 80% |
| 26-27 | 50% | 70.5672% | 80% |
| 28-29 | 100% | 70.5672% | 80% |
| 30-31 | 66.6667% | 70.5672% | 80% |
| 32-33 | 100% | 70.5672% | 80% |
| 34-35 | 50% | 70.5672% | 80% |
| 36-37 | 66.6667% | 70.5672% | 80% |
| 38-39 | 83.3333% | 70.5672% | 80% |
| 40-41 | 80% | 70.5672% | 80% |
| 42 | 100% | 70.5672% | 80% |

*The median fulfillment of the intervention indicator was 71%.*

The intervention indicator (including both offered and received interventions) was reached several times during the period, but the variation was random, since the curve with 17 eligible observations crosses the median nine times and the longest series is four observations.

**Figure S6. Implementation of intervention part (received only)**

*The median fulfillment of the Intervention indicator were 71%.*

|  | Andel | Median | Kvalitetsmål |
| --- | --- | --- | --- |
| 10-11 | 0% | 5.88235% | 80% |
| 12-13 | 0% | 5.88235% | 80% |
| 14-15 | 0% | 5.88235% | 80% |
| 16-17 | 0% | 5.88235% | 80% |
| 18-19 | 0% | 5.88235% | 80% |
| 20-21 | 0% | 5.88235% | 80% |
| 22-23 | 0% | 5.88235% | 80% |
| 24-25 | 0% | 5.88235% | 80% |
| 26-27 | 0% | 5.88235% | 80% |
| 28-29 | 0% | 5.88235% | 80% |
| 30-31 | 0% | 5.88235% | 80% |
| 32-33 | 0% | 5.88235% | 80% |
| 34-35 | 50% | 5.88235% | 80% |
| 36-37 | 33.3333% | 5.88235% | 80% |
| 38-39 | 16.6667% | 5.88235% | 80% |
| 40-41 | 0% | 5.88235% | 80% |
| 42 | 0% | 5.88235% | 80% |

*The median fulfillment of the intervention indicator was 6%.*

Exclusion of offered, but declines interventions lowered the fulfillment of the intervention indicator. The variation is non-random, as the curve crosses the median twice and the longest sequence is 12 observations.
