## Supplemental File 2 for "Translation of clinical practice guidelines on lifestyle behavior in a psychiatric setting. A discourse analytical case study"

Supplementary file 2 - Focus group interviews

The focus groups interviews were semi-structured based on pre-determined open-ended questions and discussion points

One researcher acted as moderator and handed out discussion cards, printed clinical practice guidelines (CPGs) and the local action plans during the focus group interviews in order to facilitate a discussion of the clinicians’ perception of the CPGs. The other researcher noted down gesticulations and notable body language. The researchers aimed to establish a permissive environment and explained firmly that the purpose of the study was not to control whether the clinicians knew or followed the CPGs, but to learn how they experience and articulate the CPGs and lifestyle prevention.

Focus group interviews were audio-recorded and transcribed into text by the researchers.

Interview-guide

### Introduction round

The moderator, the co-moderator and the participants all start by giving a short presentation of their names, professions etc.

### Interview guide

| **Main “task”** | **Support questions** | **Attention** |
| --- | --- | --- |
| ***Introductory – descriptive questions*** | | |
| Imagine you must explain a colleague from another specialty, how you, at your department, focus on lifestyle risk factors: diet, smoking, alcohol and exercise. | Can you try to describe a typical course of the lifestyle program? | Is the lifestyle program articulated as something they must do or something they just do? Is it prioritized?  How does habitus appear? |
| ***Theme: Needs/prioritisation*** | | |
| **Card:** “Physical health among patients with psychiatric diseases”  **Card:** “Prevention of lifestyle risk factors”  **Card:** “Prioritization of the lifestyle program” | What are your thoughts about the high mortality in psychiatry due to somatic diseases?  Discuss which patients you think need lifestyle risk factors prevention. | Which verbs are used according to how serious the problem (poor lifestyle) is?  Primary, secondary, or tertiary prevention?  Which words do they use to assess how important it is to prevent lifestyle risk factors? |
| ***Theme: The clinicians’ ownership*** | | |
| How can you clinicians play a part in prevention of lifestyle risk factors among the patients?  **Hand out the action plan –** free discussion | Discuss how you think the lifestyle program affect the patients’ lifestyles in the long run.  What is the background of this program? | Do the clinicians take ownership and appreciate the program?  Are there parts of the lifestyle program they express more ownership to than others?  Do the clinicians know about the action plan, and do they express ownership of it?  How are demands from the CPG developers/politicians translated?  What discourses are drawn on? |
| ***Theme: Implementation*** | | |
| Discuss how far you are with the implementation of the lifestyle program.  **Card:** Extracts from the National Danish Survey of Patient Experiences. Discuss the results.  **Final question**  Do you have any proposals for improvements of the lifestyle program? | Is documentation routine practice?  Why do you think that only few patients seem to experience lifestyle prevention during hospitalization? | Focus on evidence, quality of life, quality indicators etc.  Do the clinicians find the existing parts of the lifestyle program relevant? |

**Cards**

| **Physical health among patients with psychiatric diseases** | **Prioritization of the lifestyle program** |
| --- | --- |
| **Prevention of lifestyle risk factors** | **Extracts from the National Danish Survey of Patient Experiences:**  **38% do not experience that clinicians have given information about the impact of lifestyle on psychiatric diseases.**  **43% have not talked about problems with their physical health.** |
